# Longitudinal analysis of COVID-19 patients shows age-associated T cell changes independent of ongoing ill-health

**DOI:** 10.1101/2020.12.17.20248401

**Authors:** Liam Townsend, Adam H Dyer, Aifric Naughton, Rachel Kiersey, Dean Holden, Mary Gardiner, Joanne Dowds, Kate O’Brien, Ciaran Bannan, Parthiban Nadarajan, Jean Dunne, Ignacio Martin-Loeches, Padraic G Fallon, Colm Bergin, Cliona O’Farrelly, Cliona Ni Cheallaigh, Nollaig M Bourke, Niall Conlon

## Abstract

The trajectory of immunological and inflammatory changes following acute COVID-19 infection are unclear. We investigate immunological changes in convalescent COVID-19 and interrogate their potential relationships with persistent symptoms, termed *long COVID*.

We performed paired immunophenotyping at initial SARS-CoV-2 infection and convalescence (n=40, median 68 days) and validated findings in 71 further patients at median 101 days convalescence. Results were compared to 40 pre-pandemic controls. Fatigue and exercise tolerance were assessed and investigated their relationship with convalescent results.

We demonstrate persistent expansion of intermediate monocytes, effector CD8+, activated CD4+ and CD8+ T cells, and reduced naïve CD4+ and CD8+ T cells at 68 days, with activated CD8+ T cells remaining increased at 101 days. Patients >60 years also demonstrate reduced naïve CD4+ and CD8+ T cells and expanded activated CD4+ T cells at 101 days. Ill-health, fatigue, and reduced exercise tolerance were common but were not associated with immunological changes.

**Graphical Abstract:** 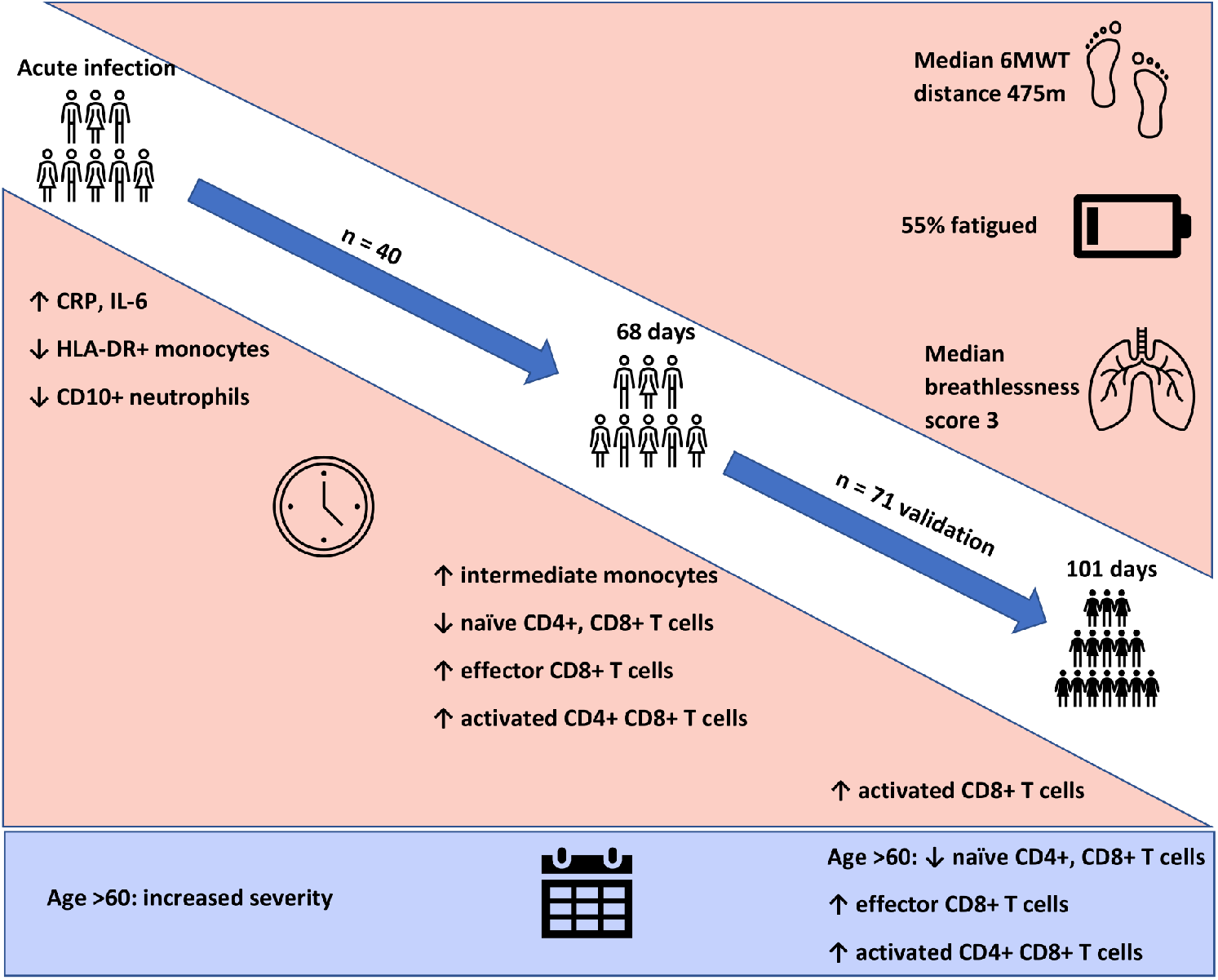

## Introduction

COVID-19, caused by the SARS-CoV-2 virus, is responsible for the largest global pandemic in modern medicine (1). The features of acute illness are well-described, ranging from disturbance in smell and mild coryzal symptoms to acute respiratory failure and the need for invasive mechanical ventilation (2, 3). Age is strongly associated with disease severity, with older individuals suffering poorer outcomes (4, 5). The immunological changes associated with severe disease are also known, with increased inflammatory proteins, coagulopathy and changes in myeloid cell populations reported (6, 7). In particular, severe COVID-19 is characterised by expansion of immature myeloid populations, with loss of HLA-DR expression by monocytes and loss of CD10 expression on neutrophils (8, 9). Panlymphopenia is also prominent, with CD4+ T cells particularly affected (10, 11).

In contrast to the well-characterised inflammatory and immunological signature of acute disease, relatively little is known about resolution of inflammatory markers and immune cell population changes during the convalescent period. These gaps in current knowledge of convalescence are of immediate importance, with the emergence of prolonged symptoms following resolution of acute infection, termed *long COVID* (12). The immunological features of this syndrome are only being described at present, with short-term follow up (less than one month) of non-hospitalised patients showing expansion of activated CD4+ and CD8+ T lymphocytes (13). Despite the importance of this issue towards understanding the long-term consequences of COVID-19, further insight into how long such changes persist, and the contribution of immune responses to the symptoms of *long COVID*, is lacking.

The clinical characteristics of long COVID are protean and an agreed definition has yet to be found; the most common symptoms include fatigue, shortness of breath and reduced exercise tolerance (14, 15). The possible mechanisms for post-COVID fatigue and breathlessness have been speculated to be associated with deconditioning, as well as hypothesised to be due to persistent inflammation or immune activation (16). However, these connections remain unexplored.

We hypothesised that, given the strong association between the immune response and severity of acute COVID-19 infection, there may be persistent and chronic changes to the immune system which may be linked to the long-term effects of COVID-19. Our goal was to determine if persistent inflammatory and immune cell dysregulation was evident in the aftermath of SARS-CoV-2 infection. We further investigated the factors that might be associated with potential persistent immune dysregulation and to interrogate the relationship between these measures and physical ill-health post-COVID-19.

## Results

### Participant characteristics

Clinic appointments were offered to 356 patients, of whom 111 (31%) attended (**Supplemental Figure 1**). We recruited three cohorts for this study. Cohort one comprised of forty participants (aged 51.4 ± 16.9 years, 52.5% female) recruited for matched longitudinal blood sampling and immunophenotyping at (i) time of initial COVID-19 disease and (ii) ten-week follow-up (median: 68, 61-72 days). Of these, 8 had mild (not hospitalised), 24 moderate (hospitalised) and 8 severe (required admission to Intensive Care Unit) acute COVID-19 disease. Cohort two comprised of 71 individuals (aged 44.3 ± 14.1 years; 70% female) recruited from the post-COVID clinic at fourteen weeks (median: 101, 76-117 days) after initial COVID-19 illness. Cohort two was younger, had lower levels of frailty and had a greater proportion of females in comparison to cohort one. Cohort two was also predominantly healthcare workers. In cohort two, 47 had mild, 18 had moderate, and 6 had severe acute COVID. All patients had lymphoid and myeloid immunophenotyping and detailed clinical and health assessments performed at outpatient appointment. The combined sample consists of both cohort one and cohort two. Detailed characteristics of cohort one, cohort two and the combined samples are presented in **Table 1**. Cohort three comprised of forty healthy pre-pandemic controls (aged 47.3 ± 15.3 years; 55% female) and was used for comparison of extended T cell immunophenotyping parameters, with 20 of these also having myeloid immunophenotyping performed. There were no age/sex differences between controls and cohort one (t = 0.9, p = 0.35; χ^2^ = 2.5, p = 0.11 respectively) or cohort two (t = 1.1, p = 0.27; χ^2^ = 0.05, p = 0.82 respectively).

**Table 1:**
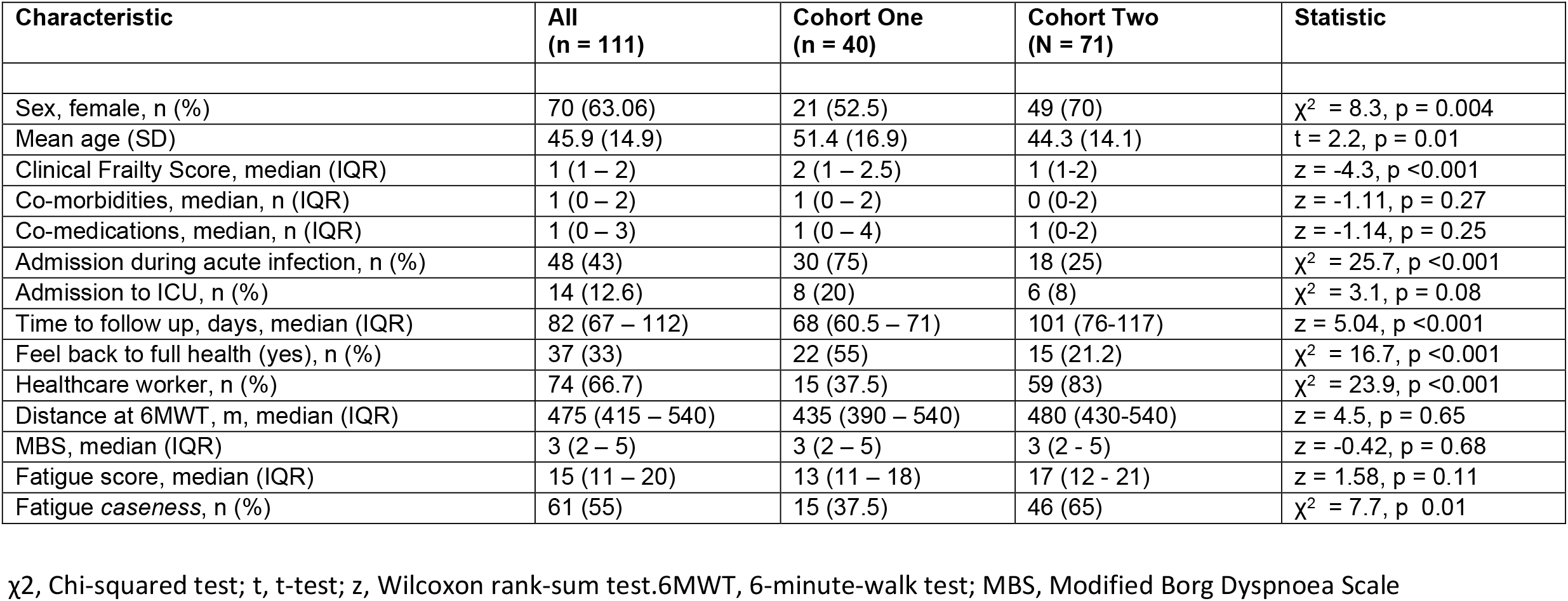
Cohort characteristics.

**Figure 1:**
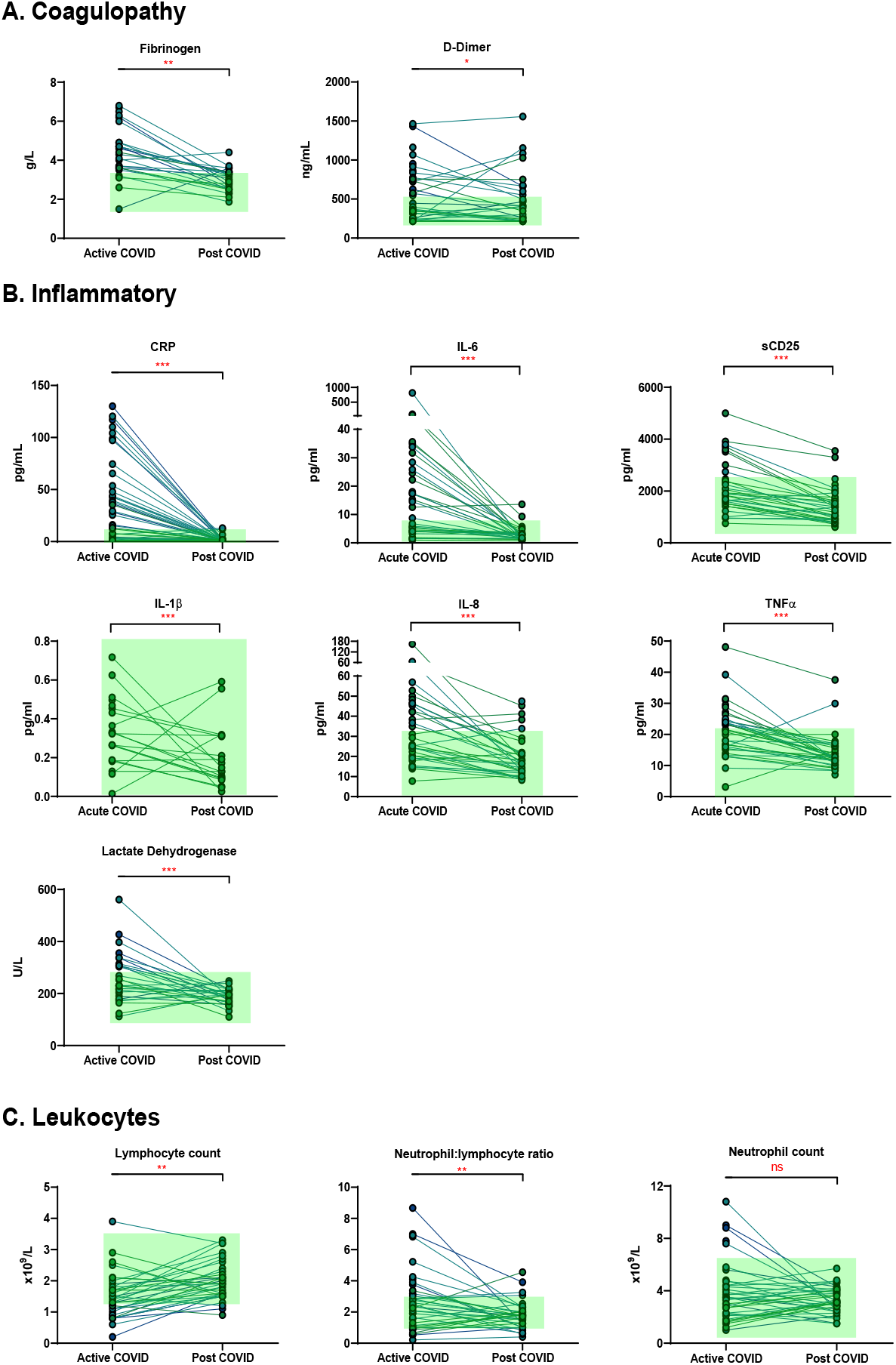
Matched results of n=40 patients from acute to convalescent (median 68 days) for: **(A)** Coagulopathy (fibrinogen, D-dimer) **(B)** Inflammatory (CRP, IL-6, sCD25, IL-1β, IL-8, TNFα, LDH) **(C)** Cell turnover (lymphocytes, neutrophil: lymphocyte ratio, neutrophils). Shaded areas show normal ranges for each measure. Differences assessed by Wilcoxon signed-rank test. * p <0.05, ** p <0.01, ***p<0.001, NS = Not Significant

### Persistent ill-health evident at 82 days following COVID-19

We first investigated the prevalence of the cardinal features of *long COVID* in our cohort. All participants were assessed for ongoing ill-health, fatigue and exercise tolerance at time of convalescent immunophenotyping (median 82 days, IQR 67 – 112). Most patients (71/111, 64%) reported that they did not feel back to full health, while 61 (55%) met the case definition for fatigue. The median fatigue score for the cohort as a whole was 15 (IQR 11 – 20), while the median fatigue score of those who met the case definition for fatigue was 20 (IQR 17 – 23) (**Table 1**). Two-thirds of participants (76/111; 68.4%) underwent a six-minute-walk test (6MWT). The median distance covered was 475m (IQR 415 – 540). The median maximal Modified Borg Dyspnoea Scale (MBS) score reported was 3 (IQR 2 – 5). The median distance covered by convalescent individuals was lower than that seen in healthy populations, but higher than that reported in post-ARDS patients (17, 18). These findings demonstrate that the primary features of *long COVID* are common in our convalescent cohort who attended for follow up appointment. We therefore wanted to further investigate if there were further physiological changes, particularly in relation to immunity, in COVID convalescence.

### Recovery from COVID-19 is associated with resolution of inflammation, coagulopathy, and cell turnover

We examined the levels of inflammatory, cell turnover and coagulation markers, all of which are known to be profoundly disturbed during acute COVID-19. Individuals in cohort one with acute COVID-19 had coagulopathy (with increased D-Dimer and fibrinogen), a marked pro-inflammatory response (elevated CRP, IL-6, TNF-α, IL-8 and IL-1β) and lymphopenia, with an increase in the neutrophil: lymphocyte ratio. All of these parameters had significantly improved by ten weeks (**Table 2** and **Figure 1**) in the majority of individuals. Several participants had persistent lymphopenia (5/40, 12.5%) and elevated D-dimer levels (10/40; 25%) at 10 weeks post infection.

**Table 2.**
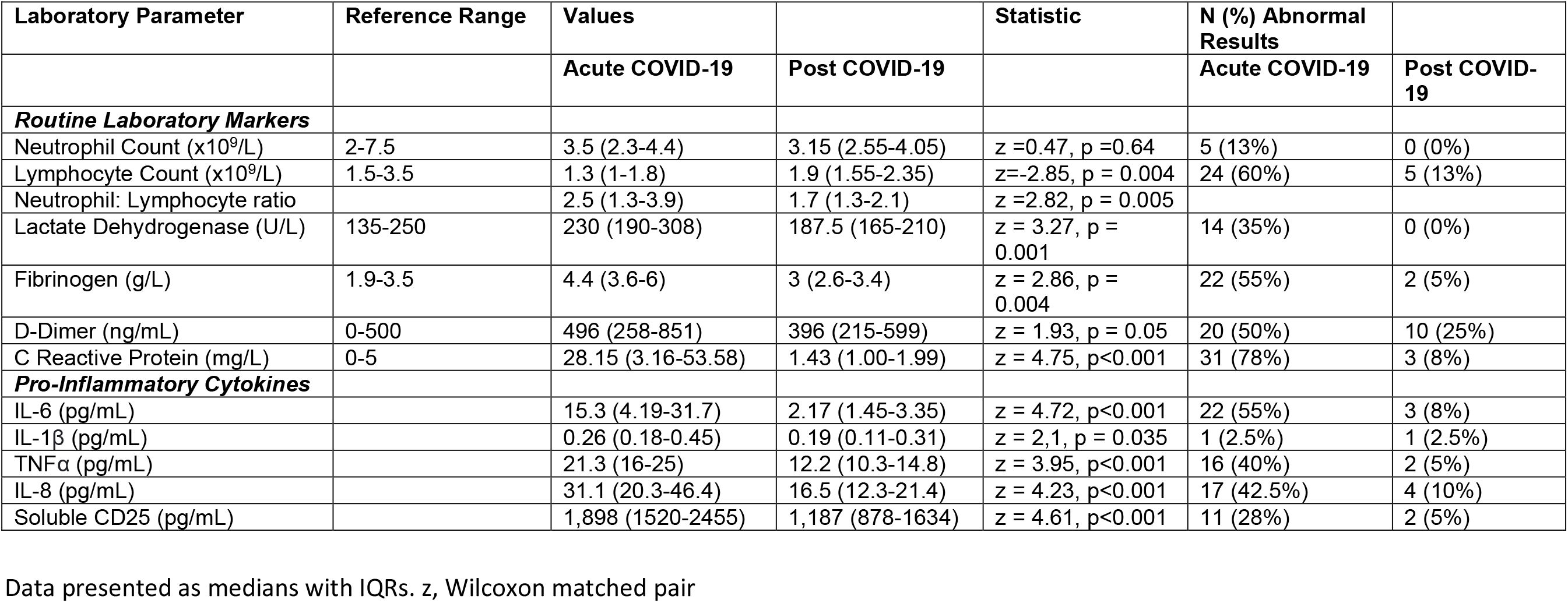
Active COVID-19 is associated with a significant pro-inflammatory response which normalises following resolution of acute infection.

### Persistent expansion of intermediate monocytes at 10 weeks post infection

We found that acute COVD-19 was associated with expansion in immature neutrophils and reduced overall CD10-expressing neutrophils (**Figures 2A, 2B**). These changes in neutrophils had resolved by ten weeks to a level comparable to that of healthy controls. While HLA-DR+ monocytes increased at convalescence to levels of healthy controls, this increase was not statistically significant; this likely reflects the mixed severities of the populations (**Figure 2C**). We also noted a significant expansion in the proportion of CD14+CD16+ intermediate monocytes in acute infection, with levels remaining significantly elevated at convalescence in comparison to controls (**Figure 2E**). Changes in the proportion of other monocyte subsets resolved to a level comparable to control participants (**Figures 2D, 2F**). We found no association between convalescent intermediate monocytes and severity of initial infection (χ^2^=0.58, p=0.76).

**Figure 2:**
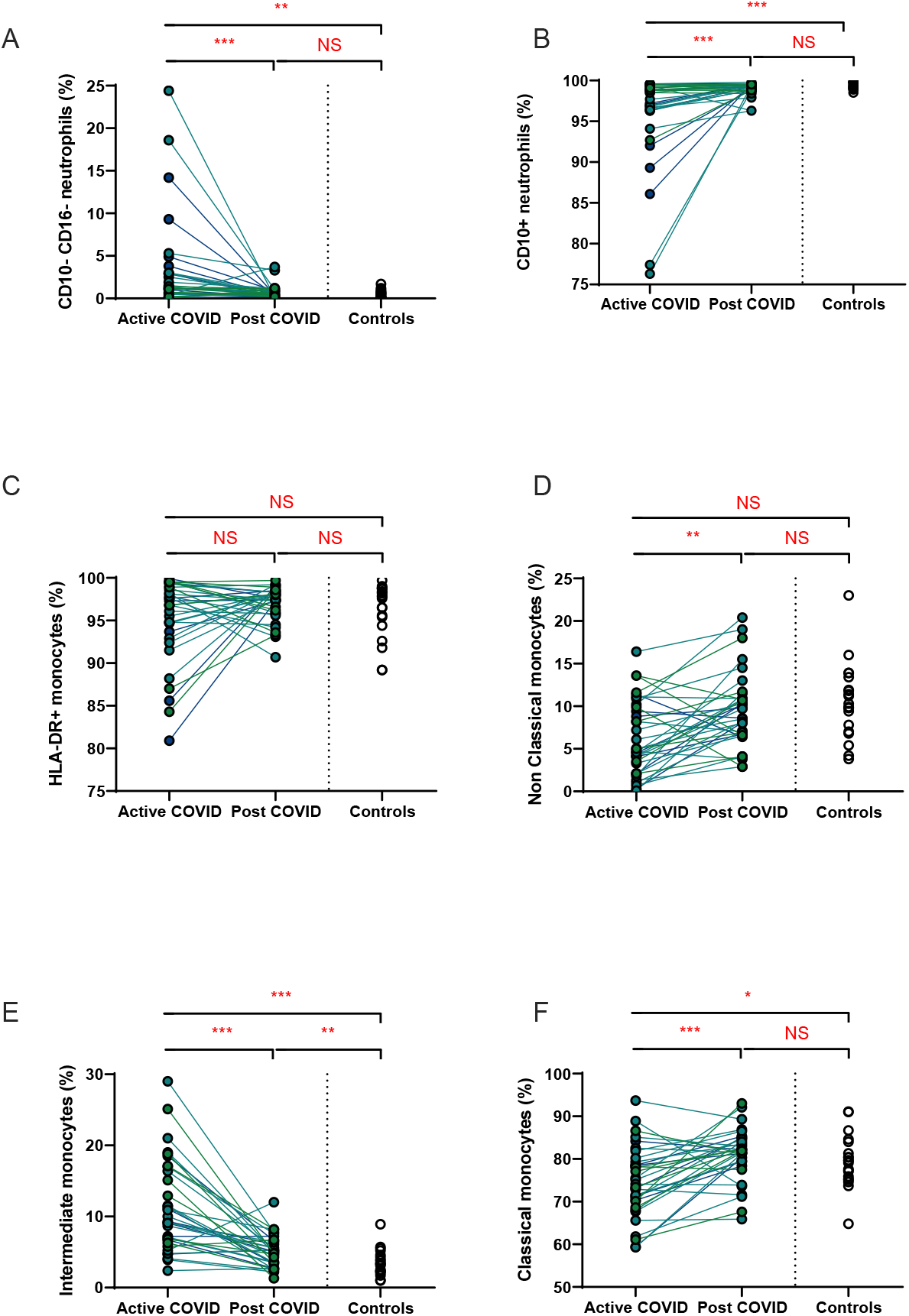
Myeloid populations from acute to convalescent COVID-19. Matched peripheral whole blood myeloid cell proportions from n=40 patients recovered from COVID-19 versus uninfected controls (n=20). **(A)** CD10-CD16-immature neutrophils **(B)** CD10+ neutrophils **(C)** HLA-DR+ monocytes **(D)** non-classical monocytes **(E)** intermediate monocytes **(F)** classical monocytes. Differences assessed using Wilcoxon rank-sum (unpaired) and Wilcoxon sign-rank (paired) tests. * p <0.05, ** p <0.01, ***p<0.001, NS = Not Significant

### Persistent changes to T cells in COVID-19 convalescence

A hallmark of acute COVID-19 is profound lymphopenia, so we sought to investigate T cell phenotypes in our longitudinal acute-convalescent cohort. As expected, acute SARS-CoV-2 infection was associated with striking lymphopenia (**Figure 3B**). We found a significant reduction in the total number of immune (CD45+) cells, total lymphocytes (CD3+), CD4+ T cells and CD8+ T cells in comparison to healthy controls (n=40). Despite this significant lymphopenia during acute COVID-19 infection, at ten weeks post-COVID all of these counts had significantly recovered and were similar to those of control participants (**Figures 3A-3D**).

**Figure 3:**
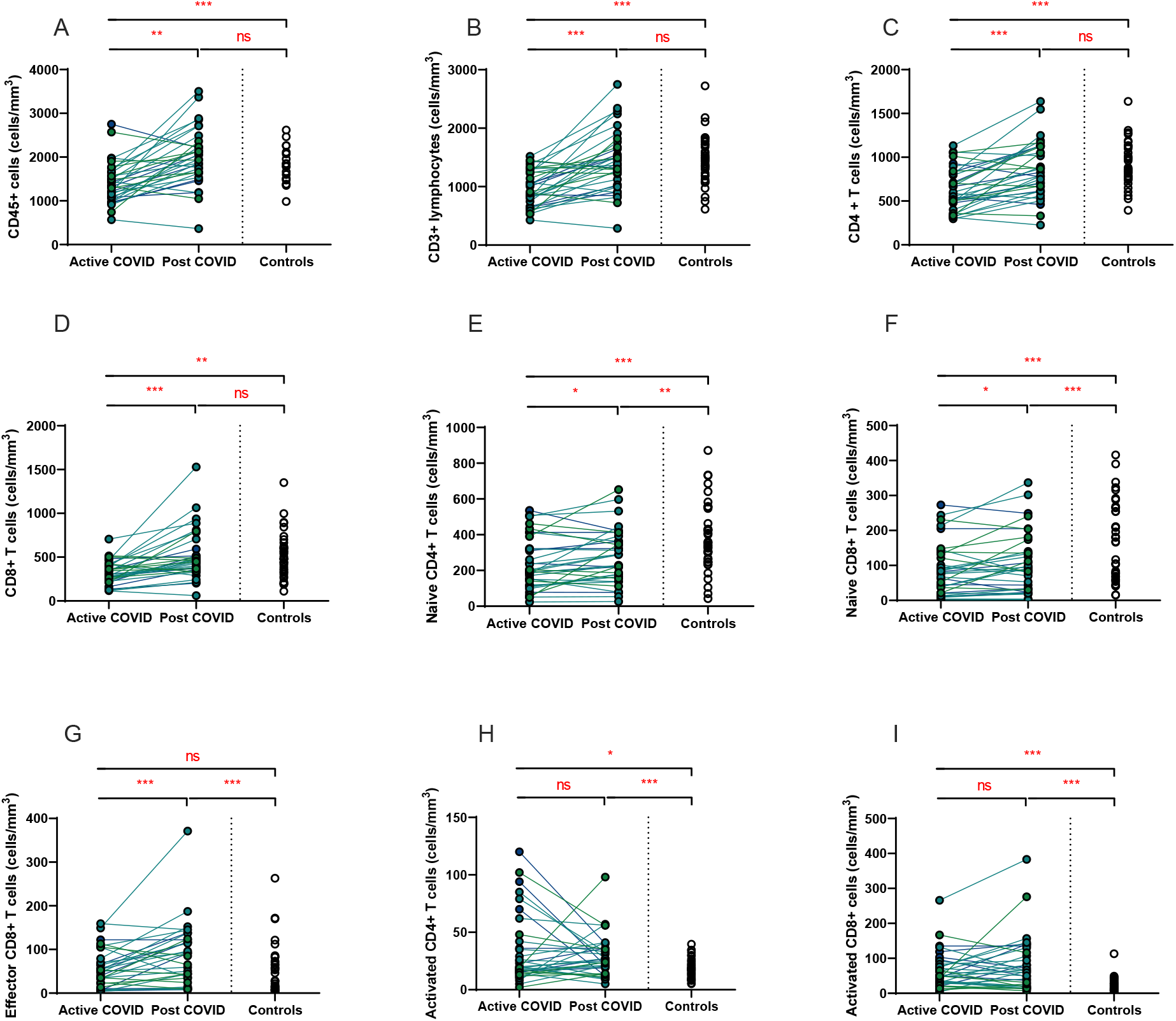
Lymphocyte subsets from acute to convalescent COVID-19. Matched peripheral whole blood lymphoid cell counts from n=40 patients recovered from COVID-19 versus uninfected controls (n=40). **(A)** CD45+ immune cells **(B)** CD3+ lymphocytes **(C)** CD4+ T cells **(D)** CD8+ T cells **(E)** naïve CD4+ T cells **(F)** naïve CD8+ T cells **(G)** effector CD8+ T cells **(H)** activated CD4+ T cells **(I)** activated CD8+ T cells. Differences assessed using Wilcoxon rank-sum (unpaired) and Wilcoxon sign-rank (paired) tests. * p <0.05, ** p <0.01, ***p<0.001, NS = Not Significant

On more detailed T cell immunophenotyping, we found that acute infection was associated with a significant reduction in absolute naïve CD4+ and CD8+ T cell counts and increased activated CD4+ and CD8+ T cell numbers in comparison to controls. Over the course of ten weeks, whilst naïve CD4+ and CD8+ cell counts had partially recovered, they remained significantly lower than the levels of healthy controls (**Figures 3E, 3F**). Numbers of effector CD8+ T cells remained expanded over the course of COVID-19 recovery and counts were significantly greater than healthy controls at ten-week follow-up (**Figure 3G**). One of the most notable differences was that activated T cell numbers did not significantly change with resolution of illness; there were significantly higher numbers of activated CD4+ and CD8+ T cells in patients ten weeks post-COVID compared to healthy controls (**Figures 3H, 3I**). Interestingly, there was no association between convalescent lymphocyte subset counts and severity of initial infection (naïve CD4+ T cells χ^2^=0.31, p=0.86, naïve CD8+ T cells χ^2^=4.12, p=0.13, effector CD8+ T cells χ^2^=0.29, p=0.19, activated CD4+ T cells χ^2^=0.28, p=0.87, activated CD8+ T cells χ^2^=0.29, p=0.07).

Alterations in the B cell compartment were relatively minor, with B cell counts recovering entirely at convalescence (**Supplemental Figure 2A**). Similarly, while acute COVID-19 was also associated with a significant decrease in NK cells, these had resolved to levels of healthy controls by ten weeks after acute infection (**Supplemental Figure 2B**). Collectively, longitudinal follow up of cohort one during convalescence revealed that while the overall panlymphopenia resolved, infected individuals maintained aberrations in certain lymphocyte subset compartments; specifically, persistently reduced naïve CD4+ and CD8+ T cells, in addition to expanded effector CD8+ T cells and activated CD4+ and CD8+ T cells at a median of 68 days following infection.

Given the persistent T-cell subset changes seen in cohort one, as well as the expansion in intermediate monocytes, we sought to validate these changes in a larger cohort of convalescent individuals with blood samples obtained at a longer follow time of 101 days (cohort two). The observations in cohort one were confirmed, with no differences in numbers of total immune (CD45+) cells, total lymphocytes (CD3+), CD4+ T cells or CD8+ T cells at 14 weeks in comparison to controls (**Supplemental Figure 3**). In contrast to the paired analysis at 10 weeks following acute infection, we observed no significant differences in naïve CD4+ or CD8+ cells, suggesting these cells had resolved to normal levels by 14 weeks in convalescence (**Figure 4**). However, the increased numbers and proportions of activated CD8+ T cells in post-COVID samples persisted, even at this longer time point of follow up in comparison to cohort one (median 101 days versus 68 days) (**Figure 4)**. There were no changes in activated CD4+ T cells seen, while convalescent COVID patients had an increased proportion of effector CD8+ T cells. The significant differences in proportion of intermediate monocytes were not seen in this cohort, with all monocyte populations returning to levels similar to controls (**Supplemental Figure 4**).

**Figure 4:**
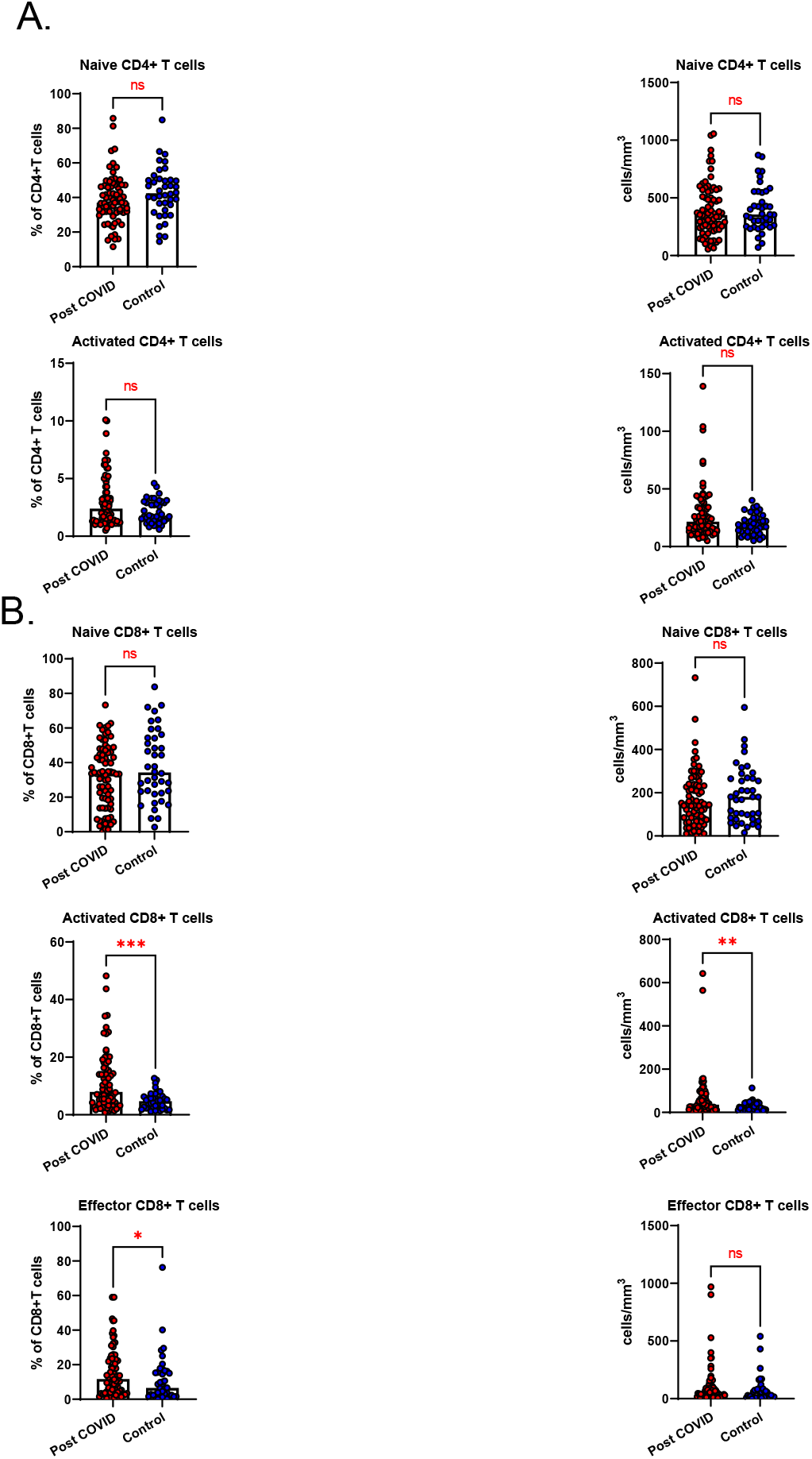
Lymphocyte subsets in convalescent COVID-19. Convalescent lymphocyte subsets from n=71 patients recovered from COVID-19 versus n=40 uninfected controls. **(A)** CD4+ T cells, showing proportion and absolute number of naïve and activated CD4+ T cells **(B)** CD8+ T cells, showing proportion and absolute number of naïve, activated and effector CD8+ T cells, Wilcoxon rank-sum test used to assess differences. * p <0.05, ** p <0.01, ***p<0.001, NS = Not Significant

Interestingly, further interrogation of these convalescent T cell results revealed that there was a relationship between these T cell changes and the persistent elevation in D-Dimer levels that we noted in our earlier analysis. Twenty patients (20/111, 18%) had an elevated D-dimer at time of assessment. D-dimer levels were negatively associated with the proportion of naive CD8+ T cells (**Figure 5A**), while they were positively associated with activated CD4+ T cells (**Figure 5B**) and activated CD8+ T cells (**Figure 5C**). However, there was no relationship between D-dimer levels and naïve CD4+ T cells (*r*^*2*^=-0.15, p=0.13) or effector CD8+ T cells (*r*^*2*^*=*0.12, p=0.24).

**Figure 5:**
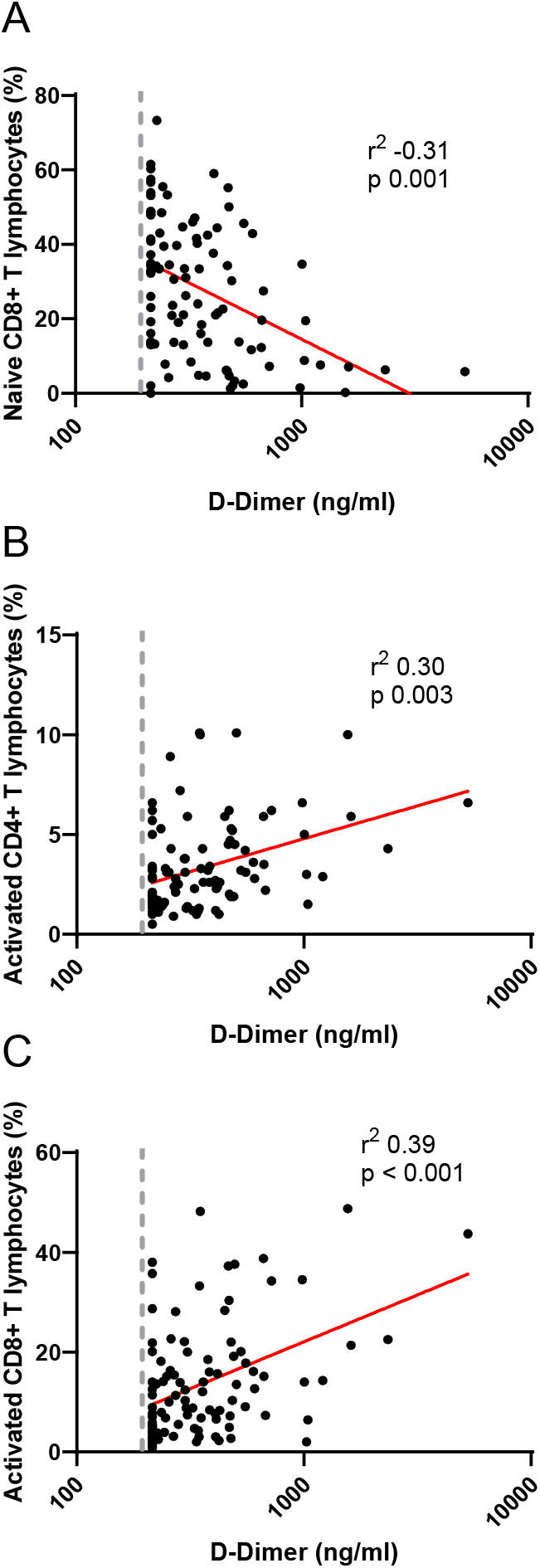
Relationship between D-dimers and lymphocyte subsets in convalescent COVID-19. Correlation between D-dimers and **(A)** naïve CD8+ T cells, **(B)** activated CD4+ T cells and **(C)** activated CD8+ lymphocytes. D-dimer shown as log scale. Dashed vertical line represents lower limit of detection for D-dimer assay. Correlation assessed by Pearson’s chi-squared test.

### Lymphocyte subset changes in convalescent COVID-19 are associated with increasing age

Older patients have worse outcomes in acute SARS-CoV-2 infection, but little is known regarding recovery of their immune system following infection. As we had noted interesting lymphocyte abnormalities post-COVID, we further investigated if age influences any of the dynamic changes T cell responses seen in convalescence. We stratified our convalescent patients into 20-year age brackets: 20 – 39 (n=43), 40 – 59 (n=49) and 60 – 80 (n=19). There was no difference in time to follow up between the three age groups (z=1.5, p=0.47). The older 60 – 80 cohort were more likely to have been admitted during acute infection than the youngest (z=4.62, p=<0.001) and 40 – 59 (z=2.77, p=0.02) cohorts. We also stratified our healthy cohort into identical age brackets: 20 – 39 (n=16), 40 – 59 (n=10), and 60 – 80 (n=14). There were distinct age-associated changes seen in lymphoid populations in the convalescent cohort, with an age-associated decline in both absolute number and proportions of naïve CD4+ and CD8+ cells and an age-associated increase in activated CD4+ and CD8+ cells (**Supplemental Figure 5**). There were minor increases in effector CD8+ T cells with age (**Supplemental Figure 5**). We also investigated disease-associated effects by comparing these results to age-matched controls. There were distinct differences noted, with the most dramatic being in the oldest cohort, with post-COVID individuals having reduced number of naïve CD4+ and naïve CD8+ (**Figures 6A, 6C**) and increased number and proportion of activated CD4+ and CD8+ cells (**Figures 6B, 6E**) compared to age-matched controls. There were no differences between the age-matched controls and infected individuals in the youngest cohort. The 40 – 59 age group showed increased number of effector CD8+ T cells (**Figure 6C**), activated CD4+ and activated CD8+ T cells in comparison to uninfected controls, with no differences in naïve T cell counts. These changes were mirrored in the proportion of T cell populations across age groups (**Supplemental Figure 6**). These data suggest that in those >60 years of age the post-COVID changes in T cells persist longer than in younger individuals.

**Figure 6:**
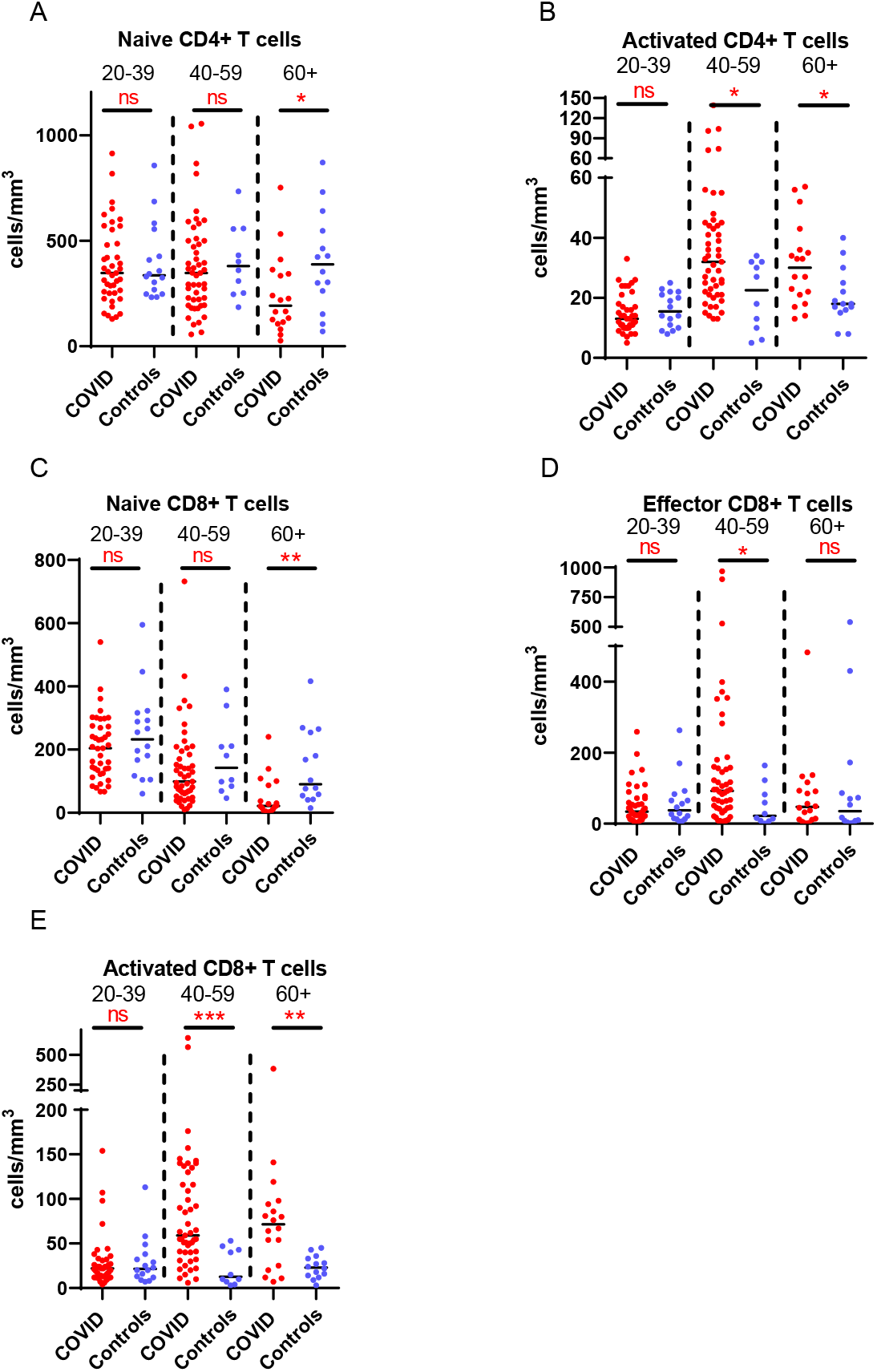
Age-associated changes in convalescent lymphocyte subsets versus age-matched controls. Lymphocyte immunophenotyping of convalescent COVID patients (n=111) broken down by age with age-matched controls, showing absolute number of **(A)** naïve CD4+ T cells and **(B)** activated CD4+ T cells, and **(C)** naïve CD8+ T cells, **(D)** activated CD8+ T cells and **(E)** effector CD8+ T cells. Differences assessed by Wilcoxon rank-sum test. * p <0.05, ** p <0.01, ***p<0.001, NS = Not Significant

### Persistent symptoms are independent of T cell immunophenotype following COVID-19 illness

We have demonstrated the high prevalence of ill-health and the cardinal features of *long COVID* in our cohort Our analysis has also revealed persistent changes to T cells, most notably activated CD8+ T cells, in the convalescent period following COVID-19. We finally wanted to investigate the relationship between immunophenotyping parameters and subjective symptoms of fatigue, as assessed by CFQ-11 score, and exercise tolerance, as assessed by performance on the 6MWT. We used linear regression under unadjusted conditions and controlled for age, sex and clinical frailty score in order to determine relationships. There were no associations between naïve CD4+, naïve CD8+, effector CD8+, activated CD4+, or activated CD8+ T cells and fatigue score, distance reached on the 6MWT or maximal MBS reported under any of the conditions examined (**Table 3, Figure 7**). We also evaluated the association between physical health measures and classical, intermediate and non-classical monocyte populations, given the intermediate monocyte differences we saw at 68 days. There were no associations with monocyte populations (**Supplemental Figure 7**).

**Table 3:** No relationship between extended T cell subsets and symptoms of fatigue, dyspnoea or distance walked on the six-minute walk test post-COVID-19.

|  | CFQ-11 |  |  |  | 6MWT<br>(meters) |  |  |  | Borg Dyspnoea<br>Scale |  |  |  |
| --- | --- | --- | --- | --- | --- | --- | --- | --- | --- | --- | --- | --- |
|  | Model 1 |  | Model 2 (adj.) |  | Model 1 |  | Model 2 (adj.) |  | Model 1 |  | Model 2<br>(adj.) |  |
| | $\beta$ Coeff<br>(95% CI) | P | Adj. $\beta$ Coeff<br>(95% CI) | p | $\beta$ Coeff<br>(95% CI) | p | Adj. $\beta$ Coeff<br>(95% CI) | p | $\beta$ Coeff<br>(95% CI) | p | Adj. $\beta$ Coeff<br>(95% CI) | p |
| **Naïve**<br>**CD4**<br>**Count** | 0.005<br>(0.00, 0.01) | 0.046 | 0.004<br>(-0.001, 0.01) | 0.07 | -0.05<br>(-0.19, 0.09) | 0.50 | -0.14<br>(-0.26, -0.02) | 0.02 | -0.0002<br>(-0.003, 0.002) | 0.83 | -0.0002<br>(-0.003, 0.002) | 0.89 |
| **Naïve**<br>**CD8**<br>**Count** | 0.01<br>(0.001, 0.02) | 0.04 | 0.01<br>(0.001, 0.02) | 0.04 | 0.20<br>(-0.03, 0.42) | 0.08 | -0.10<br>(-0.34, 0.13) | 0.39 | 0.001<br>(-0.003, 0.005) | 0.70 | 0.003<br>(-0.002, 0.008) | 0.21 |
| **Effector**<br>**CD8**<br>**Count** | -0.002<br>(-0.01, 0.01) | 0.63 | 0.003<br>(-0.05, 0.05) | 0.92 | -0.04<br>(-0.20, 0.12) | 0.65 | 0.01<br>(-0.13, 0.15) | 0.85 | -0.002<br>(-0.005, 0.001) | 0.22 | -0.002<br>(-0.004, 0.001) | 0.23 |
| **Activated**<br>**CD4**<br>**Count** | 0.04<br>(-0.69, 0.77) | 0.82 | -0.01<br>(-0.06, 0.04) | 0.75 | -1.04<br>(-2.31, 0.23) | 0.11 | -0.0002<br>(-1.19, 1.19) | 0.99 | -0.0001<br>(-0.02, 0.02) | 0.99 | -0.01<br>(-0.03, 0.02) | 0.48 |
| **Activated**<br>**CD8**<br>**Count** | -0.01<br>(-0.02, 0.01) | 0.32 | -0.01<br>(-0.02, 0.004) | 0.20 | -0.08<br>(-0.35, 0.19) | 0.56 | 0.09<br>(-0.16, 0.33) | 0.48 | -0.003<br>(-0.008, 0.002) | 0.23 | -0.004<br>(-0.01, 0.001) | 0.10 |

**Figure 7:**
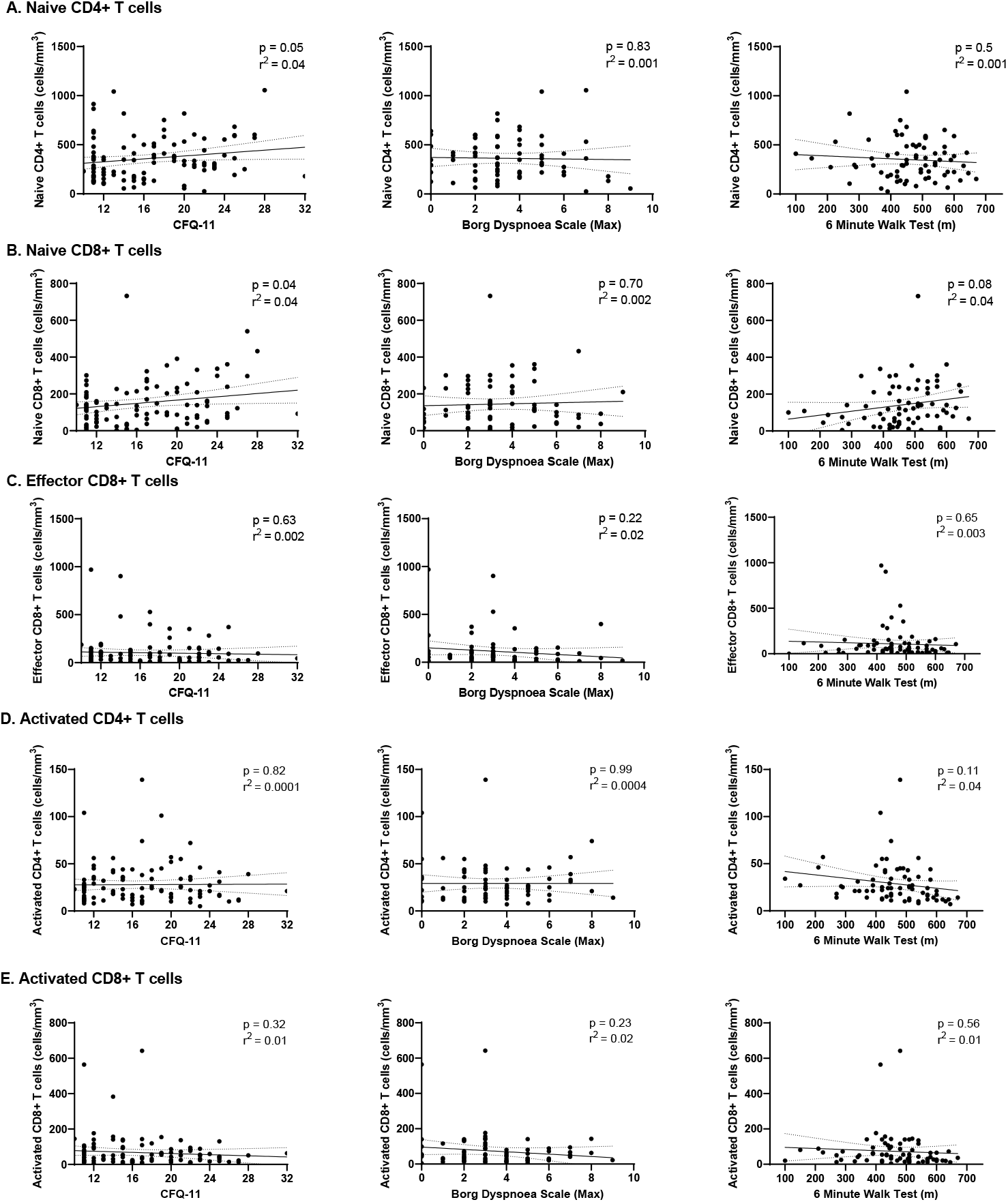
Relationships of lymphocyte subsets with physical health measures and D-dimers. Relationship with fatigue, perceived exertion and 6MWT distance with **(A)** naïve CD4+ T cells, **(B)** naïve CD8+ T cells, **(C)** effector CD8+ T cells, **(D)** activated CD4+ T cells and **(E)** activated CD8+ T cells. Relationships assessed using linear regression. CFQ-11 = Chalder Fatigue Questionnaire-11

These findings suggest that persistent ill-health and fatigue are frequently reported at a medium-term interval following COVID-19, but they are independent of any persistent changes to the immune parameters investigated in this study.

## Discussion

We show evidence of persistent abnormalities of T cell responses in the aftermath of acute SARS-CoV-2 infection that are unrelated to initial disease severity across the spectrum of disease, with severity ranging from mild disease managed in the community to requirement for ICU care. Specifically, we demonstrate an expansion of effector CD8+ T cells, activated CD4+ and CD8+ T cell populations and reduction in naïve CD4+ and CD8+ T cells at ten weeks following acute infection. Further analysis of COVID convalescence found that the expansion of activated CD8+ T lymphocytes is still evident at a median of 101 days following infection. Equally notable is the recovery post-COVID of myeloid populations to levels similar to healthy controls. While we report persistent expansion of intermediate monocytes at 68 days, all monocyte populations have normalized at 101 days. Given that the myeloid compartment is significantly altered in acute infection, it is reassuring to see resolution of these changes at convalescence. Similarly, routine clinical measures of inflammation and coagulation also return to normal levels. The exception to this is D-dimers, which remain elevated in 18% of patients. Persistent ill-health is common in our cohort, with almost two-thirds (64%) not feeling back to full health, and fatigue seen in more than half the cohort. These are the cardinal features described in *long COVID*. However, these findings of persistent ill-health are independent of immunological parameters measured.

While persistent expansion of activated T lymphocytes are described in the setting of chronic viral infection, they are less commonly seen following infection by an acute pathogen (19). However, persistent activation of CD8+ T cells has been reported in the setting of parvovirus B19 infection, as well as the aftermath of severe influenza A H7N9 infection (20, 21). Interestingly, we demonstrate that D-dimer levels are closely associated with both activated CD4 and CD8 lymphocyte counts. This persistent elevation of D-dimers mirrors early reports from elsewhere (22). We now report that the elevations in D-dimers is associated with increased levels of activated T lymphocytes and reduced levels of naïve T lymphocytes. Potential mechanisms for this include persistent endothelial dysfunction following resolution of infection, as well as the degradation of extra-vascular sites of coagulation (23, 24). D-dimers have also been shown to be markers of crosstalk between the coagulation system and adaptive immune system in chronic viral infection such as HIV (25). The data we present here highlight the need for further investigations of such crosstalk in the context of COVID-19.

One of the most striking findings was that these T cell-specific changes are most marked in older >60-year-old participants, with persistent abnormalities across naïve and activated CD4+ and CD8+ T cells, while those aged <40 show complete resolution of these changes. While age is known to affect immune recovery in treated chronic infections such as HIV, there is relatively little known about ageing and immune recovery following acute viral infections (26). The ageing immune system has been implicated in the ageing-associated mortality rate in acute COVID-19 (27). T cell responses are crucial in modulating the immune response and preventing host damage in acute viral infections such as influenza (28). Impairment of the adaptive immune system with age, in particular immunosenescence, has been well-described (29). This is of particular importance when considering the impact of lasting immunity in previously infected older individuals and how little is known about adaptive immune responses in this population in the context of COVID-19. The ability to sustain an effective memory immune response to infection has been shown to diminish with age (30). A recent study found there was a general decline in recognition of viral-associated peptides between the ages of 42 and 58 in a pre-pandemic cohort (31). Another important consideration regarding our results is how such alterations may impact on the potential long-term efficacy of vaccination of older cohorts, where responses have previously been shown to be highly variable (32, 33). These results emphasize the importance of further study into activation and resolution of immune responses in SARS-CoV-2 infection in older individuals.

Interestingly, we found the cardinal features of *long COVID*, namely the presence of fatigue and reduced exercise tolerance, had no association with lymphocyte subset changes. Nonetheless, we demonstrate a large burden of fatigue, breathlessness, and ill-health in this cohort. This is reflective of previous clinical descriptors of *long COIVD* cohorts. Our group have previously reported a high prevalence of post-COVID fatigue, which was independent of severity of initial infection (34). These findings have been replicated in subsequent studies showing persistent ill-health in young, otherwise healthy individuals (35). Our findings further highlight the difficulty in establishing a reliable biomarker for ongoing ill-health following COVID-19 infection.

While it is likely that the proportion of those reporting persistent ill-health is likely confounded by their increased likelihood to attend for follow-up, it demonstrates a significant burden of morbidity. It will be illuminating to follow the prevalence of autoimmune disease in the general population in the post-pandemic era. This is particularly relevant given the concept of bystander T cell activation during viral infection. Bystander T cell recruitment has been described in both hepatitis A and influenza A infection (36, 37). While the activation of such T cells are considered beneficial in the control of acute infection and protective immunity, such non-specific immune activation has been associated with immunopathology, or host damage (38). Indeed, activation of both CD4+ and CD8+ lymphocytes in acute infection has been associated with the development of a wide array of autoimmune conditions (39, 40). The potential for SARS-CoV-2 to cause immunopathology has been proposed as a mechanism behind multisystem inflammatory syndrome in children (MIS-C), a new paediatric inflammatory condition associated with COVID-19 (41). Autoantibodies against multiple cell types have been demonstrated in this condition, while immune complex formation has also been implicated (42, 43). Furthermore, this is similar to the immunopathology seen in other post-infectious autoimmune conditions, such as Kawasaki disease (44). Pro-thrombotic autoantibody generation has also been described in acute COVID-19 in adult populations, which would link both activated T lymphocytes and elevated D-dimers (45). D-dimer levels have also been reported to correlate with CD8+ lymphocytes in MIS-C (46). Further characterisation of these persistently activated cells is warranted to inform assessment of their functional consequences.

Our study has several limitations worth noting. It is a single-centre study at a single medium-term interval. However, we have two separate convalescent time points, in addition to data from acute illness. This allows a disease and recovery trajectory to be plotted. We have loss to follow-up, with 31% of patients attending their outpatient appointments. This is a common challenge seen in research conducted in clinical ambulatory care settings. However, our cohort may have an increased burden of symptoms following COVID-19 than that seen in the entire affected population.

The results reported here provide insights into the immune consequences of SARS-CoV-2 infection, as well as the age effects on immune recovery. It provides possible mechanisms for immunopathology and should inform the design of ongoing studies into the immunological consequences of COVID-19 and associations with long COVID clinical features.

In conclusion, we report several key findings that add significant knowledge regarding resolution of the immunological responses in the convalescent period of COVID-19 infection. Encouragingly, our matched longitudinal patient data shows that all cell counts return towards levels of healthy controls. Although there are persistent lymphocyte and monocyte abnormalities at 68 days, these had resolved by 101 days post infection with the exception of a persistent expansion of activated CD8+ T cells. We show that age, while being strongly associated with poor outcome in acute COVID, is also strongly associated with impaired immunological recovery in convalescence. The association of D-dimer levels with activated lymphocytes provide a potential basis for persistent immune dysfunction following SARS-CoV-2 infection. While we demonstrate the burden of persistent ill-health following SARS-CoV-2 infection, this was not associated with immunological changes.

## Methods

### Study setting and participants

This study was carried out in the post-COVID-19 clinic at St James’s Hospital, Dublin, Ireland. Appointments were offered to all individuals with a positive SARS-CoV-2 nasopharyngeal swab PCR at our institution between March and May 2020, including both those hospitalised and those managed in the community during their acute illness. Severity of initial infection was graded as mild (did not require hospitalisation), moderate (required hospitalisation) or severe (required intensive care unit admission). Appointments were offered at a minimum of six weeks following resolution of symptoms or hospital discharge. Outpatient appointments were not offered to residents in long term care facilities.

### Inflammatory makers and immunophenotyping

Blood sampling was incorporated as part of routine phlebotomy occurring on the same day as study participation. An identical sampling and analytic pipeline was implemented on the cohort during acute infection. IL-6, IL-8, IL-1β, TNF-α and soluble CD25 levels were measured in serum by ELISA (Ella ProteinSimple). Immunophenotyping by flow cytometry was carried out on fresh whole EDTA-treated blood and samples were analysed on a FACS Canto II Flow Cytometer (BD San Jose USA), using BD DIVA v8 and FLO Jo v10 software. BD FACSCanto™ clinical software was used for acquisition of BD Multitest™ 6-colour TBNK and TruCount tubes. All other immunophenotyping samples were analysed using BD DIVA v8 and FLO Jo v10 software (**Supplemental Figure 8**). The frequency and absolute cell counts of CD45+ T cells (CD3+, CD4+ and CD8+), B cells (CD19+) and NK cells (CD16+CD56+) were generated by BD Multitest™ 6-colour TBNK and TruCount method. Naive (CD27+) and effector (CD27-) CD4+ or CD8+ T cells were characterised for expression of CD27, CD45RA and CD197. T cell activation was assessed by CD38 and HLA-DR expression. Absolute cell counts for naïve effector and activated T cells were calculated using the absolute frequencies of parent populations acquired from the BD TruCount tubes. Cell phenotyping assays were validated and accredited in line with ISO15189 standards. Classical, intermediate and non-classical monocytes were characterised by CD14 and CD16 expression. The maturation status of CD16+ neutrophils was evaluated by CD10 expression. Antibodies used in flow cytometry phenotyping are in **Supplemental Table 1**. The reference ranges for all assays were generated using a panel of 40 healthy controls and were established in a pre-pandemic setting.

### Physical Health Assessment

Physical health assessment occurred at time of outpatient assessment and convalescent immunophenotyping. Patients were asked a binary question as to whether or not they felt back to full health. Fatigue was assessed using the Chalder Fatigue Scale (CFQ-11) (47, 48). Participants answer eleven questions in relation to physical and psychological fatigue, with reference to the past month in comparison to their pre-COVID-19 baseline. A Likert scale (0-3) is used to measure responses, constructing a total score ranging from 0 to 33 (49).

The CFQ-11 also allows differentiation of “cases” vs “non-cases” where scores 0 and 1 (*Better than usual/No worse than usual*) are scored a zero and scores 2 and 3 (*Worse than usual/Much worse than usual*) are scored a 1 (bimodal scoring). Those with a total score of four or greater are considered to meet the criteria for fatigue. This latter method for *caseness* resembles other fatigue questionnaires (49-52).

To assess cardiopulmonary and musculoskeletal function, a 6MWT was used, with total distance covered recorded (53, 54). The MBS, widely used in both healthy and diseased states to analyse exertion during submaximal exercise, assessed perceived exertion during the 6MWT (range 0 - 10) (55, 56).

### Statistical analysis

All statistical analysis was carried out using STATA v15.0 (Texas, USA) and statistical significance considered p<0.05. Descriptive statistics are reported as means with standard deviations (SD) and median with interquartile ranges (IQR) as appropriate.

Between-group differences were assessed using t-tests, Wilcoxon rank-sum and chi-square tests according to underlying data type and distribution. Paired analysis of laboratory parameters and immune cell populations were carried out using Wilcoxon sign-rank test. Linear regression was used to model the relationship between immune cell parameters (independent variable) and CFQ-11/6MWT (distance covered in metres) / MBS score using separate linear models. These were performed unadjusted in the first instance (Model 1), followed by adjustment for age, sex, and severity of initial infection (Model 2). Results are presented as β coefficients with corresponding 95% confidence intervals (Cis) and *p* values. Correlation analysis between parameters was performed using Spearman correlation tests.

### Study approval

Ethical approval for the current study was obtained from the Tallaght University Hospital (TUH)/St James’s Hospital (SJH) Joint Research Ethics Committee (reference REC 2020-03). Informed consent was obtained from all participants in the current study in accordance with the Declaration of Helsinki (57).

## Data Availability

All data is available within the manuscript

## Author contributions

Conceptualisation: LT, AHD, COF, CNC, NMB and C.N.C.; Methodology: LT, AHD, AN, RK, DH, MG, JDo, KOB, PF, NMB and NC. Formal Analysis: LT, AHD, JDu and NMB Investigation: LT, AN, RK, DH, MG, JDo, KOB, CBa, PN, IML; Resources: JDu, IML, CBe, COF, CNC, NMB and NC.; Data Curation: LT, AHD, AN, RK, JDu and NC; Writing-Original Draft: LT, AHD, CNC, NMB and NC; Writing-Review & Editing: AN, JDu, IML, PF, CBe, COF, CNC, NMB and NC; Visualisation: LT, AHD, AN, RK, DH, MG and NMB; Supervision: IML, CBe, COF, CNC, NMB and NC; Funding Acquisition: LT and NC

## Acknowledgements

We would like to thank all patients involved in the study, the support provided by the Clinical Research Facility at Saint James’s Hospital, the STAR-Bioresource team, as well as the work of the clinicians and multi-disciplinary team in the outpatient clinic.

---

**Supplemental Figure 1:**
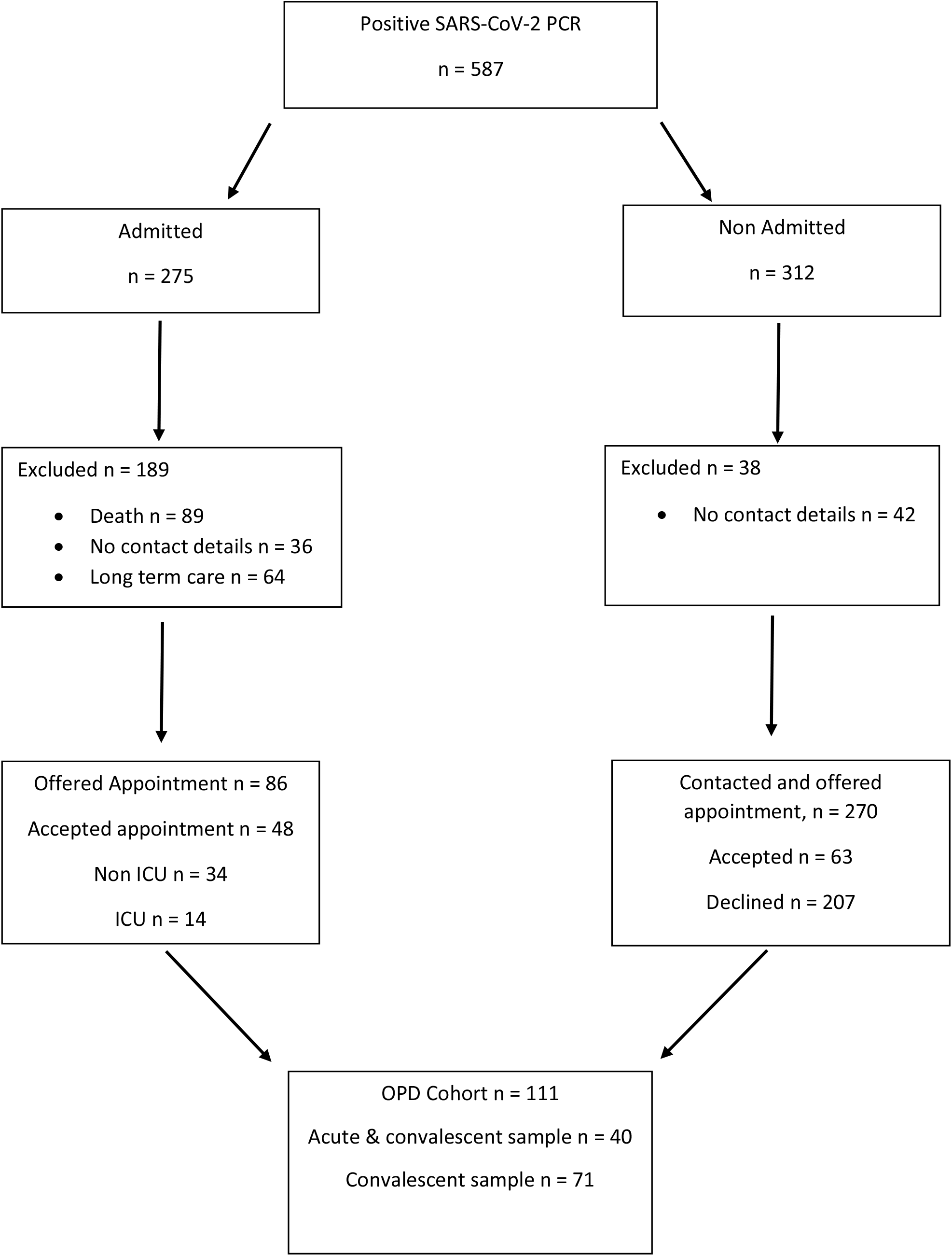
Patient enrolment diagram.

**Supplemental Figure 2:**
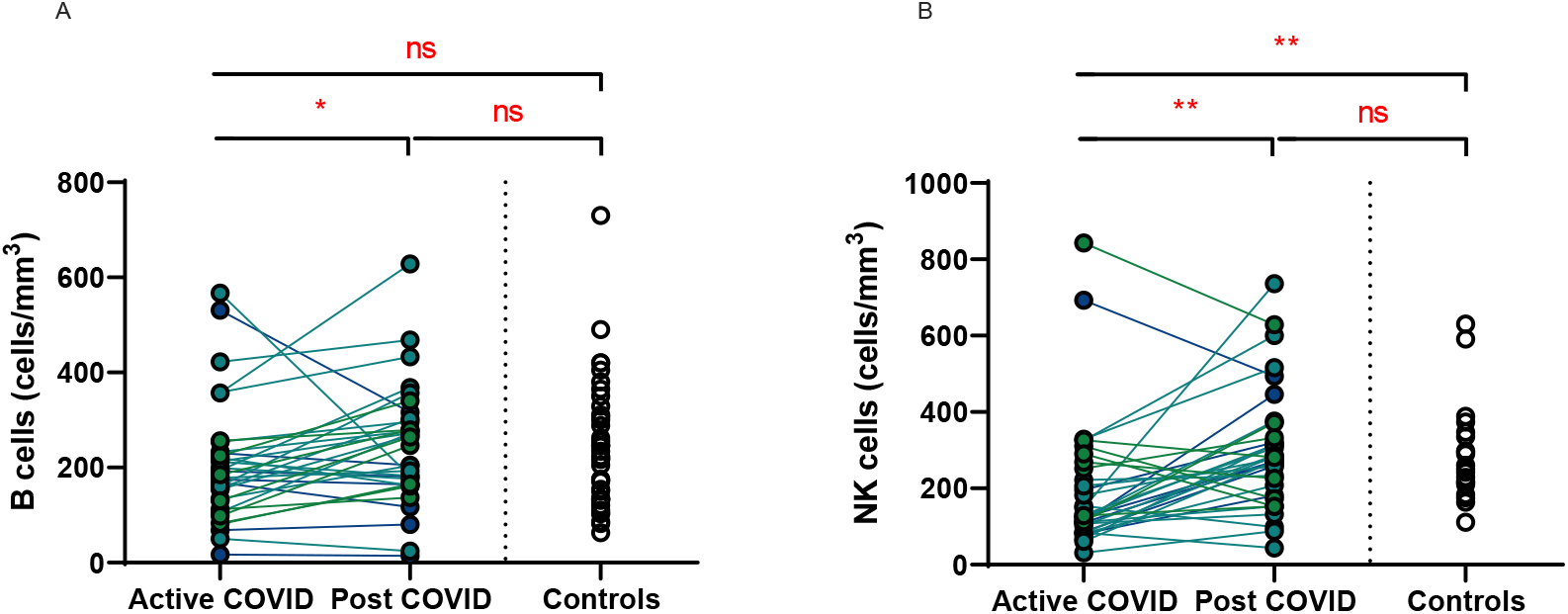
Recovery of B and NK cells. Matched peripheral whole blood lymphoid cell counts from n=40 patients recovered from COVID-19 versus uninfected controls (n=40). **(A)** B cells **(B)** NK cells. Wilcoxon rank-sum (unpaired) and Wilcoxon sign-rank (paired) tests.* p <0.05, ** p <0.01, ns = Not Significant

**Supplemental Figure 3:**
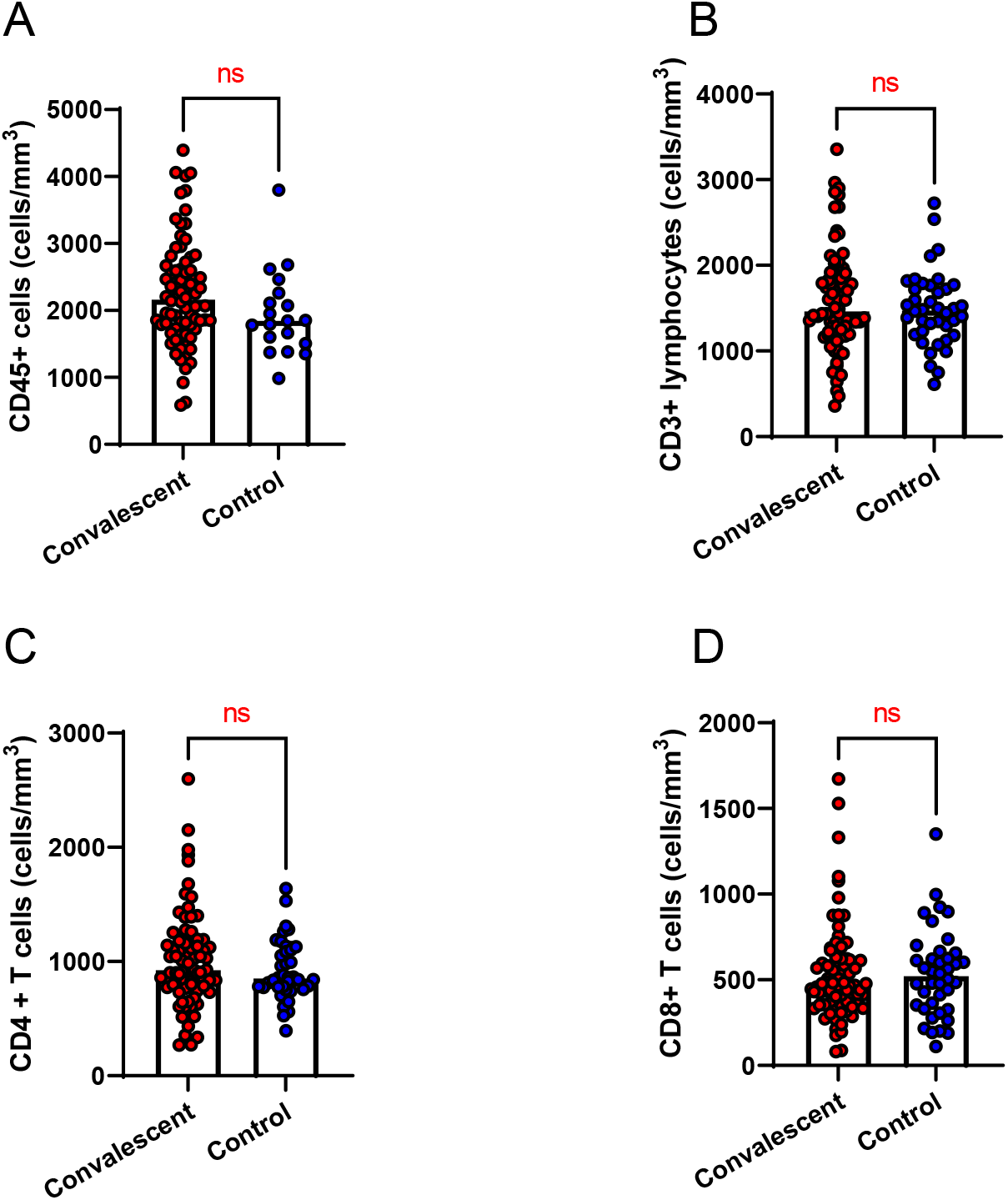
Major lymphoid populations in convalescent COVID-19. Major lymphoid populations in n=71 convalescent COVID patients in comparison to n=40 controls **(A)** CD45+ cells **(B)** CD3+ lymphocytes **(C)** CD4+ T cells **(D)** CD8+ T cells. Wilcoxon rank-sum test ns=not significant

**Supplemental Figure 4:**
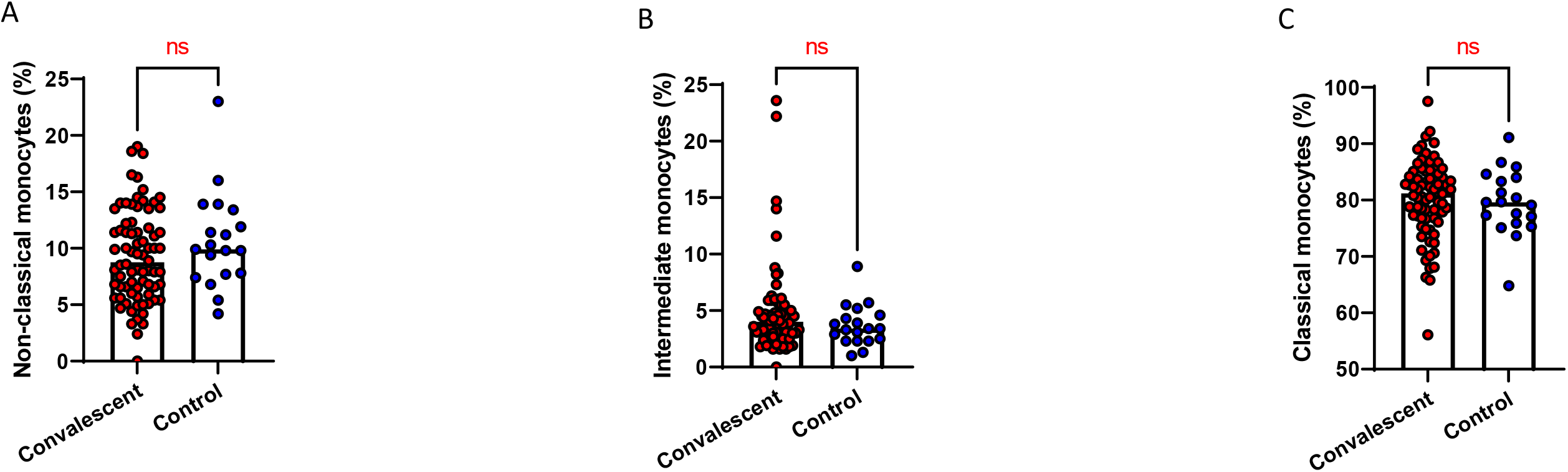
Monocyte subpopulations in convalescent COVID-19. Monocyte subpopulations in n=71 convalescent COVID patients in comparison to n=20 controls **(A)** non-classical monocytes **(B)** intermediate monocytes **(C)** classical monocytes. Wilcoxon rank-sum test. ns=not significant

**Supplemental Figure 5:**
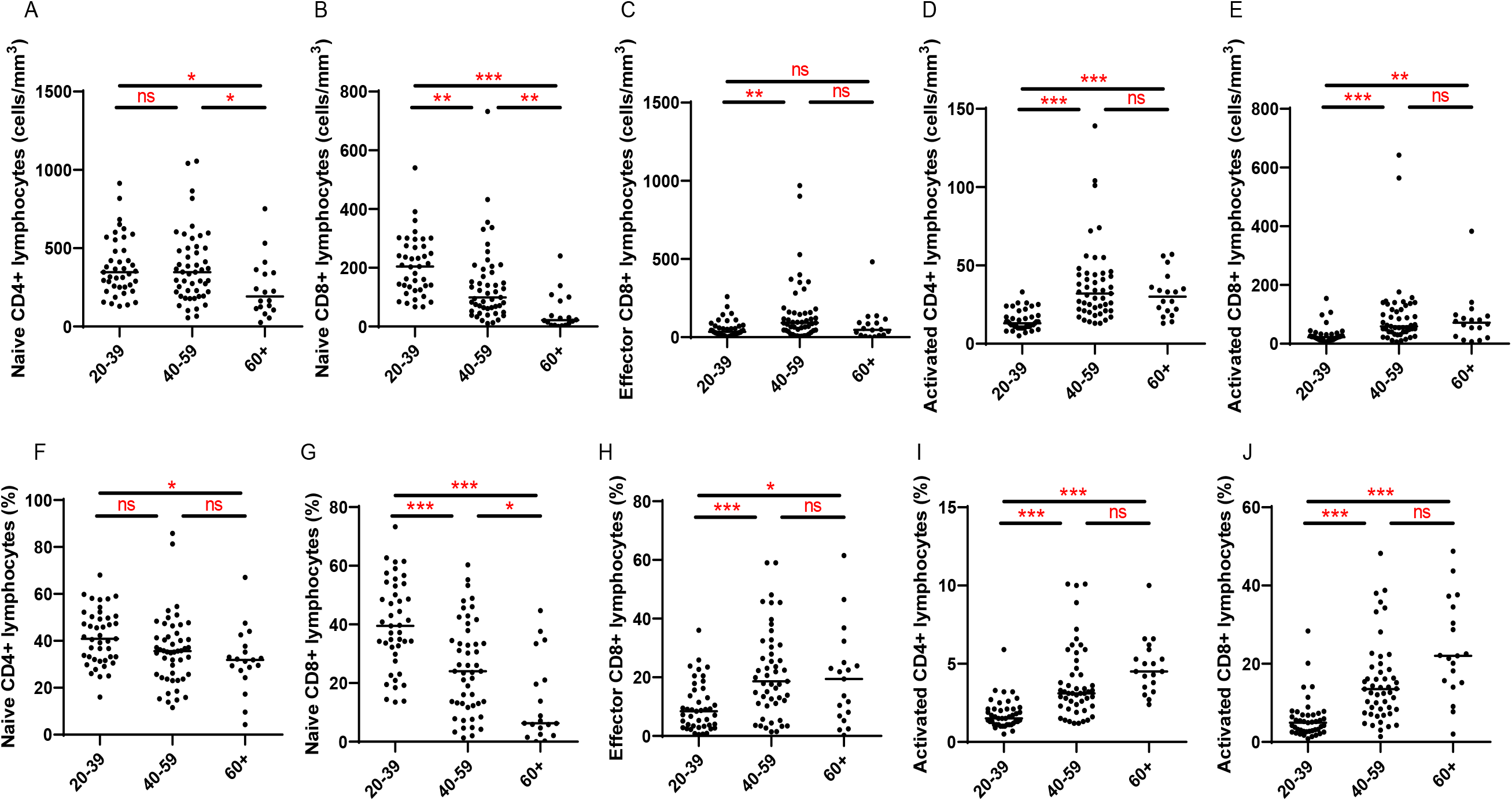
Age-associated changes in convalescent lymphocyte subsets. Lymphocyte immunophenotyping of convalescent COVID patients (n=111) broken down by age. **(A)** naïve CD4+ T cell count **(B)** naïve CD8+ T cell count **(C)** effector CD8+ T cell count **(D)** activated CD4+ T cell count **(E)** activated CD8+ T cell count **(F)** naive CD4+ T cell proportion **(G)** naïve CD8+ T cell proportion **(H)** effector CD8+ T cell proportion **(I)** activated CD4+ T cell proportion **(J)** activated CD8+ T cell proportion. Kruskal-Wallis test with post-hoc Dunn test.* p <0.05, ** p <0.01, ***p<0.001, ns = Not Significant

**Supplemental Figure 6:**
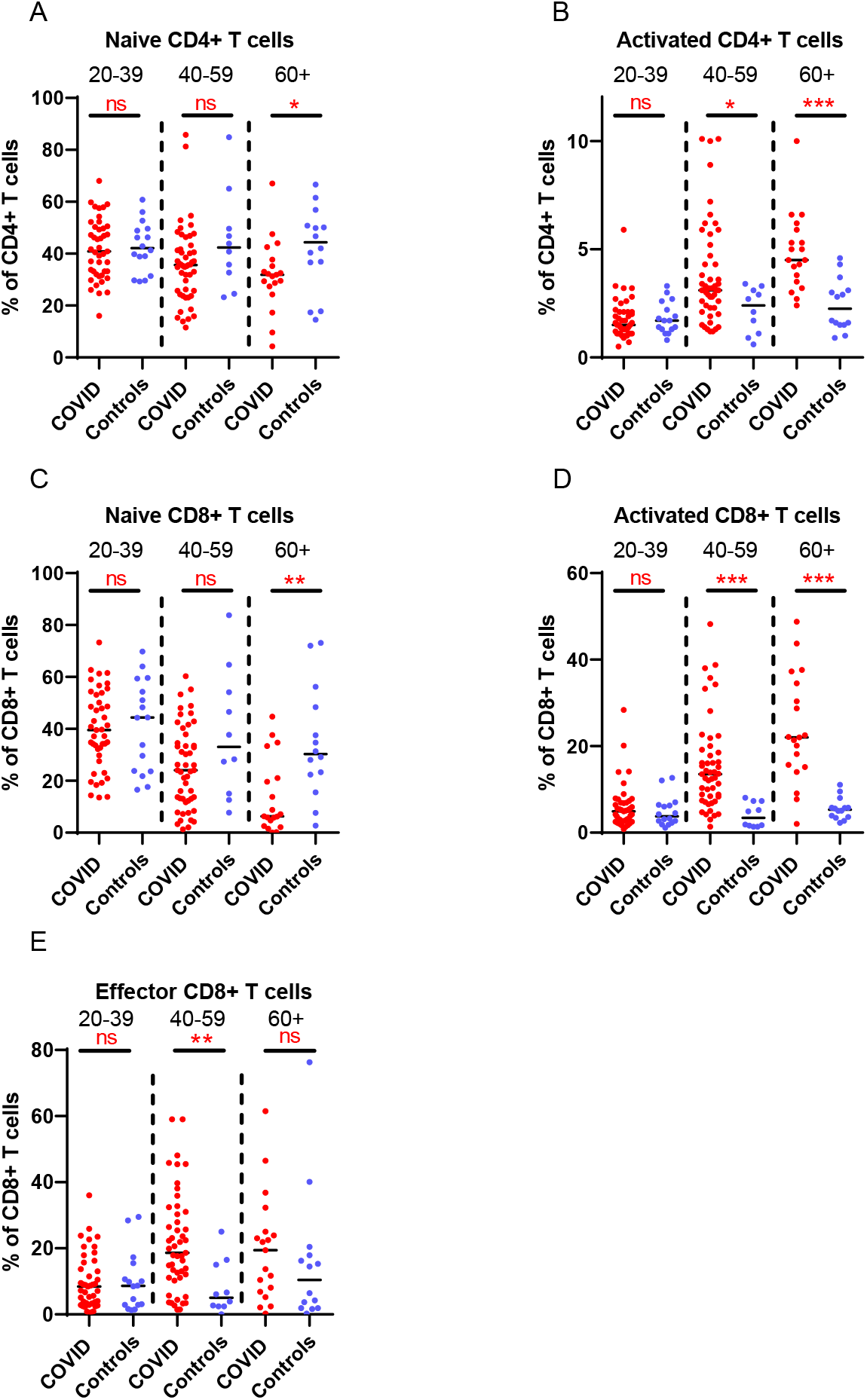
Age-associated changes in convalescent lymphocyte subset proportions versus age-matched controls. Lymphocyte immunophenotyping of convalescent COVID patients (n=111) broken down by age with age-matched controls, sowing proportions of naïve (A) and activated (B) CD4+ T cells, and naïve (C), activated (D) and effector (E) CD8+ T cells. Differences assessed by Kruskal-Wallis test with post-hoc Dunn test.* p <0.05, ** p <0.01, ***p<0.001, NS = Not Significant

**Supplemental Figure 7:**
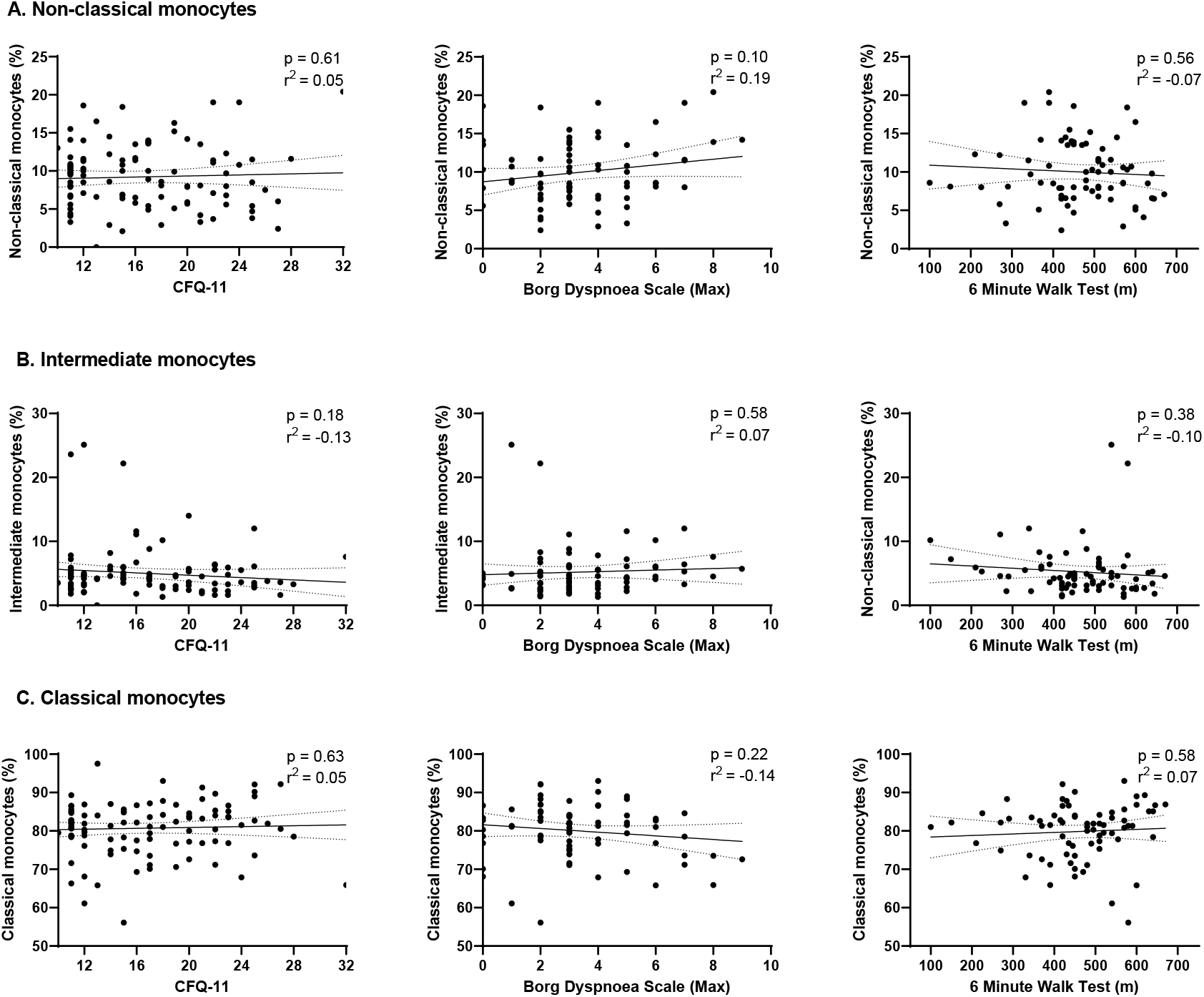
Relationships of monocyte subsets with physical health measures. Relationship with fatigue, perceived exertion and 6MWT distance in n=101 convalescent COVID patients with (A) non-classical monocytes (B) intermediate monocytes (C) classical monocytes. Correlation assessed with Pearson’s chi-squared test. CFQ-11 = Chalder Fatigue Questionnaire-11

**Supplemental Figure 8:**
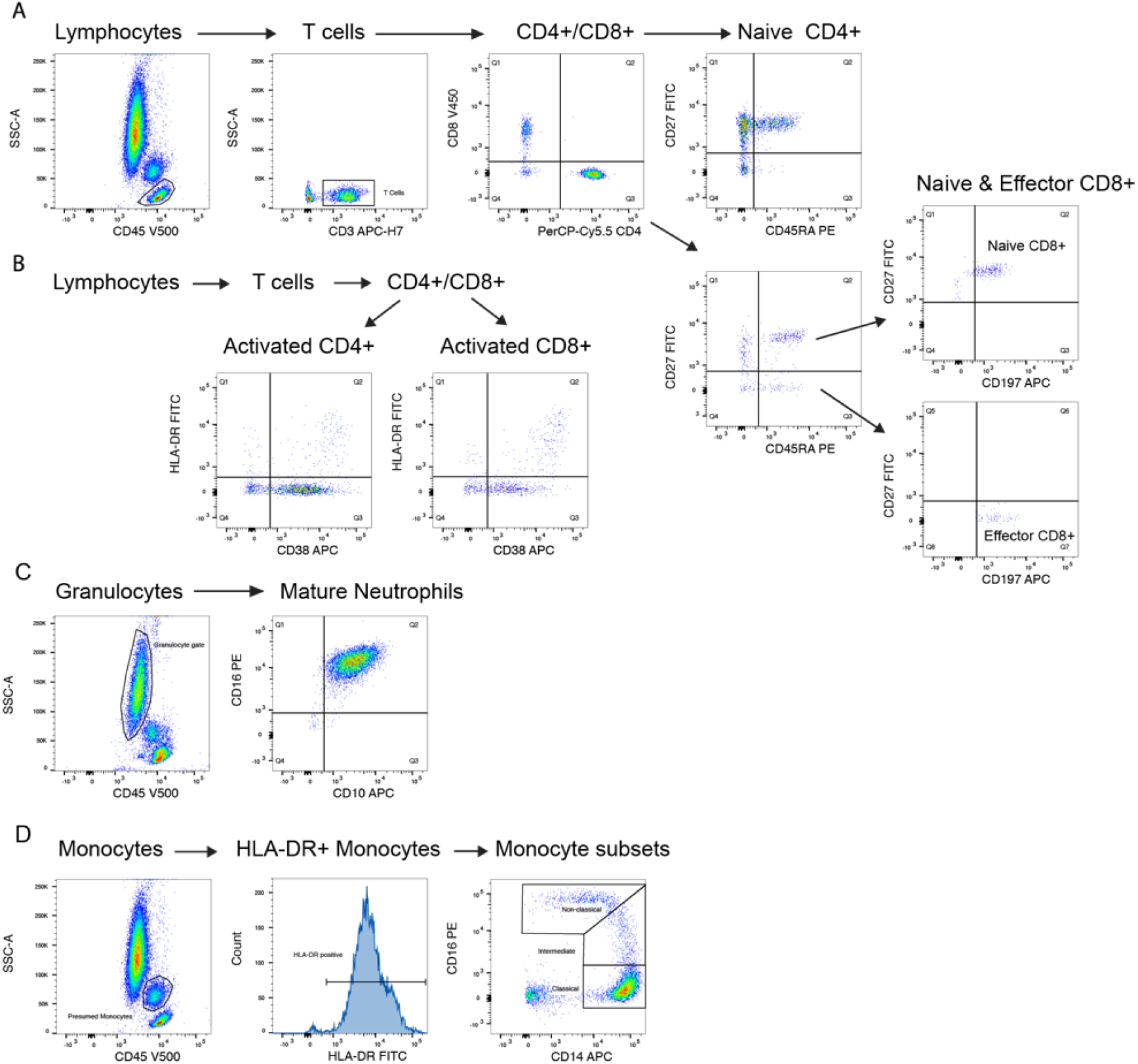
Flow cytometry gating strategy. Gating strategy for flow cytometry shown **(A)** naïve and effector T lymphocytes **(B)** activated T lymphocytes **(C)** neutrophils **(D)** monocytes

**Supplemental Table 1:** Flow cytometry antibodies.

| Antibody | Fluorophore | Clone | Company | Catalogue # |
| --- | --- | --- | --- | --- |
| CD45 | V500-C | 2D1 | BD Biosciences | 655873 |
| CD8 | V450 | RPA-T8 | BD Biosciences | 560347 |
| CD3 | APC-H7 | SK7 | BD Biosciences | 641415 |
| CD4 | PerCP/CY5.5 | Sk3 | BD Biosciences | 332772 |
| CD45RA | PE | - | BD Biosciences | 556627 |
| CD27 | FITC | - | BD Biosciences | 555440 |
| CD197 | Alexa Fluor 647 | 150503 | BD Biosciences | 560816 |
| HLA DR | FITC | L243 | BD Biosciences | 347400 |
| CD38 | APC | HB-7 | BD Biosciences | 345807 |
| CD14 | APC | MΦP9 | BD Biosciences | 345787 |
| CD16 | PE | B73.1 | BD Biosciences | 332779 |
| CD10 | APC | HI10a | BD Biosciences | 332777 |

